# SARS-CoV-2 IgG seroprevalence surveys in blood donors before the vaccination campaign, France 2020-2021

**DOI:** 10.1101/2022.07.29.22278190

**Authors:** Pierre Gallian, Nathanaël Hozé, Nadège Brisbarre, Paola Mariela Saba Villarroel, Elif Nurtop, Christine Isnard, Boris Pastorino, Pascale Richard, Pascal Morel, Simon Cauchemez, Xavier de Lamballerie

## Abstract

We conducted a cross-sectional study for SARS-CoV-2 anti-S1 IgG prevalence in French blood donors (n=32605), from May-2020 to January-2021. A mathematical model combined seroprevalence with daily number of hospital admissions to estimate the probability of hospitalization upon infection and determine the number of infections while correcting for antibody decay. There was an overall seroprevalence increase over the study period and we estimate that ∼15% of the French population had been infected by SARS-CoV-2 by January-2021. The infection/hospitalization ratio increased with age, from 0.56% (18-30yo) to 6.75% (61-70yo). Half of the IgG-S1 positive individuals had no detectable antibodies 4 to 5 months after infection. The seroprevalence in group O donors (7.43%) was lower (p=0.003) than in A, B and AB donors (8.90%). We conclude, based on seroprevalence data and mathematical modelling, that the overall immunity in the French population before the vaccination campaign started was too low to achieve herd immunity.

## INTRODUCTION

By mid-April 2022, the pandemic caused by SARS-CoV-2 has been responsible for more than 6.2 million deaths worldwide and more than 500 million people have been diagnosed with COVID-19. In France, since January 2020, over 27 million cases and 144,000 deaths have been recorded (COVID-19 Dashboard by the Center for Systems Science and Engineering at Johns Hopkins University). The pandemic continues to grow and hopes for curbing the dynamics of infections rely heavily on building up population immunity derived from natural infection or vaccines. Estimating the proportion of the population carrying antibodies is necessary to ascertain the degree of viral penetration, estimate the level of protection against infection and ascertain disease severity. Seroprevalence surveys in the blood donor population provide rapid, large-scale information on humoral immunity that can be used to map affected areas and monitor epidemic progression over time. Serological studies on SARS-CoV-2 are technically challenging but important given the proportion of asymptomatic forms (Mizumoto et al., 2020). Amongst patients, antibody levels appear to be higher and more persistent in those with a severe form of the infection compared to those with a moderate form. Reinfections are possible mainly in people with low levels of neutralizing antibodies (Jeffery-Smith et al., 2021). Finally, anti-SARS-CoV-2 antibodies decrease or disappear over time and these kinetics differ according to the immunoglobulin and antigenic target sought (Pelleau et al., 2021).

We report here a study of the seroprevalence of anti-SARS-CoV-2 IgG antibodies in the French blood donor population (called Covidonneur) before the implementation of the general population vaccination campaign and before the circulation of antigenic variants. Using a mathematical model, we analyze these data jointly with age-stratified hospital admissions data to reconstruct epidemic dynamics in France.

## RESULTS

### SARS-CoV-2 anti-S1 IgG prevalence

A total of 32,605 donations (17,043 females and 15,562 males) from 31,922 donors (16,758 females (52.5%) and 15,164 males (47.5%)) were collected between the end of March 2020 and early February 2021 during six collection times over four survey’s periods chosen according to the evolution of the COVID-19 epidemic in France (Table 1, Figure S1). SARS-CoV-2 anti-S1 IgG prevalence rates for each department according to the sampling period and evolution over time are shown in Figure 1. In January/February 2021 (survey time: T6), no significant differences in rates of anti SARS-CoV-2 IgG according to gender could be found (Table 1 and Figure 2). Seroprevalence was highest (p<0.001) in the 18-30 age group (10.90%; 95%CI 9.90-11.90) and lowest among the 31-40 (6.66%; 95%CI 5.60-7.72, p=0.004) and 61-70 (5.46%; 95%CI 4.27-6.65, p<0.001) age groups compared to other age groups (Figure 2). The time trends of the seroprevalence rate varied with age (Figure 3A). In blood donors over 40 years of age, seroprevalence rates ranged from 2.66% (95%CI 1.41-3.91) to 8.67% (95%CI 6.57-10.77) during the study. In the age group 31-40 years old, there was a decrease in seroprevalence over time and in the age group 18-30 years old seroprevalence increased from the summer of 2020.

**Table 1:**
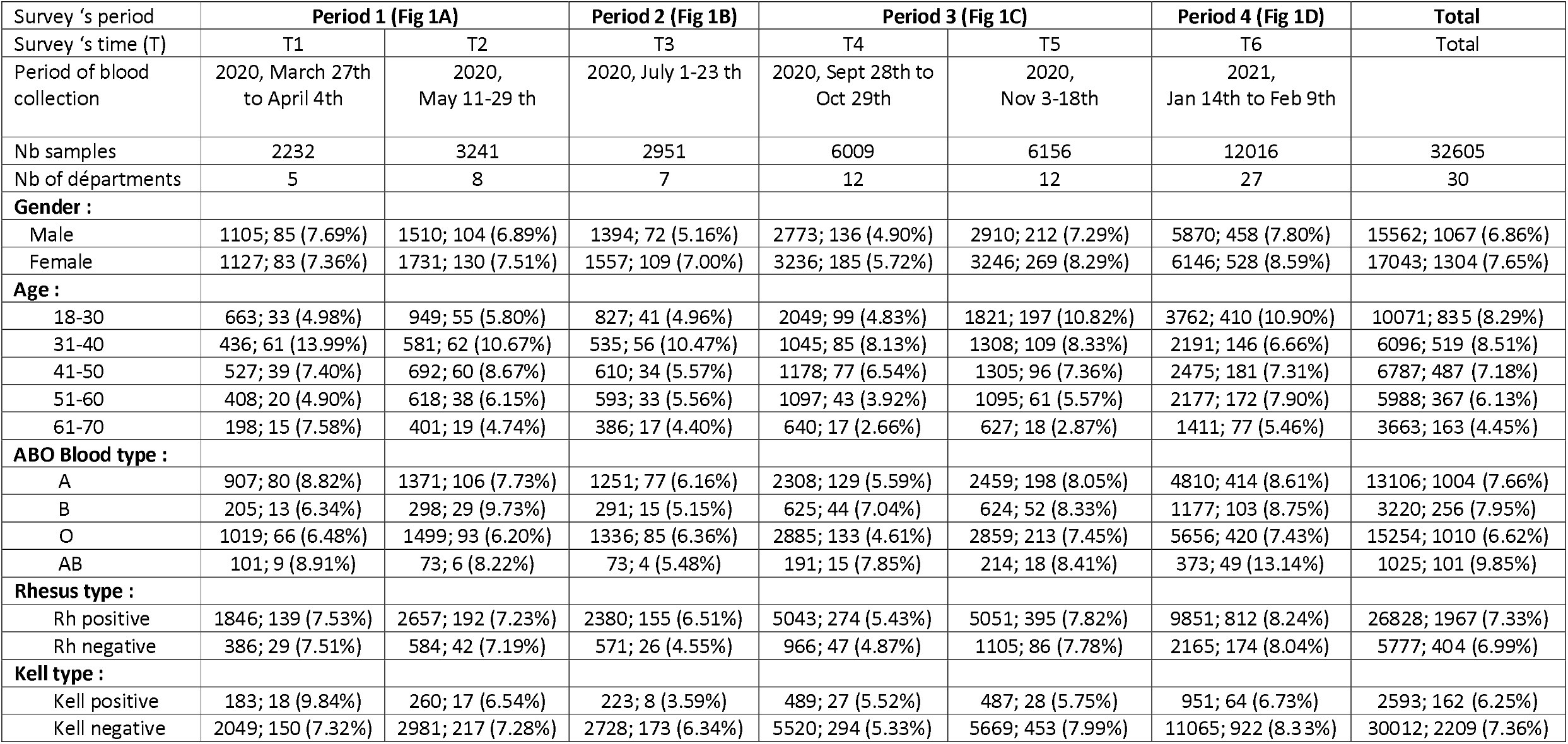
Anti-SARS-CoV-2 prevalence by demographic characteristics for the 4 period of survey and detailed for the 6 survey’s times. [n; pos (%age)]

| Survey 's period | Period 1 (Fig 1A) |  | Period 2 (Fig 1B) | Period 3 (Fig 1C) |  | Period 4 (Fig 1D) | Total |
| --- | --- | --- | --- | --- | --- | --- | --- |
| Survey 's time (T) | T1 | T2 | T3 | T4 | T5 | T6 | Total |
| Period of blood collection | 2020, March 27th to April 4th | 2020, May 11-29 th | 2020, July 1-23 th | 2020, Sept 28th to Oct 29th | 2020, Nov 3-18th | 2021, Jan 14th to Feb 9th |  |
| Nb samples | 2232 | 3241 | 2951 | 6009 | 6156 | 12016 | 32605 |
| Nb of départements | 5 | 8 | 7 | 12 | 12 | 27 | 30 |
| Gender : |  |  |  |  |  |  |  |
| Male | 1105; 85 (7.69%) | 1510; 104 (6.89%) | 1394; 72 (5.16%) | 2773; 136 (4.90%) | 2910; 212 (7.29%) | 5870; 458 (7.80%) | 15562; 1067 (6.86%) |
| Female | 1127; 83 (7.36%) | 1731; 130 (7.51%) | 1557; 109 (7.00%) | 3236; 185 (5.72%) | 3246; 269 (8.29%) | 6146; 528 (8.59%) | 17043; 1304 (7.65%) |
| Age : |  |  |  |  |  |  |  |
| 18-30 | 663; 33 (4.98%) | 949; 55 (5.80%) | 827; 41 (4.96%) | 2049; 99 (4.83%) | 1821; 197 (10.82%) | 3762; 410 (10.90%) | 10071; 835 (8.29%) |
| 31-40 | 436; 61 (13.99%) | 581; 62 (10.67%) | 535; 56 (10.47%) | 1045; 85 (8.13%) | 1308; 109 (8.33%) | 2191; 146 (6.66%) | 6096; 519 (8.51%) |
| 41-50 | 527; 39 (7.40%) | 692; 60 (8.67%) | 610; 34 (5.57%) | 1178; 77 (6.54%) | 1305; 96 (7.36%) | 2475; 181 (7.31%) | 6787; 487 (7.18%) |
| 51-60 | 408; 20 (4.90%) | 618; 38 (6.15%) | 593; 33 (5.56%) | 1097; 43 (3.92%) | 1095; 61 (5.57%) | 2177; 172 (7.90%) | 5988; 367 (6.13%) |
| 61-70 | 198; 15 (7.58%) | 401; 19 (4.74%) | 386; 17 (4.40%) | 640; 17 (2.66%) | 627; 18 (2.87%) | 1411; 77 (5.46%) | 3663; 163 (4.45%) |
| ABO Blood type : |  |  |  |  |  |  |  |
| A | 907; 80 (8.82%) | 1371; 106 (7.73%) | 1251; 77 (6.16%) | 2308; 129 (5.59%) | 2459; 198 (8.05%) | 4810; 414 (8.61%) | 13106; 1004 (7.66%) |
| B | 205; 13 (6.34%) | 298; 29 (9.73%) | 291; 15 (5.15%) | 625; 44 (7.04%) | 624; 52 (8.33%) | 1177; 103 (8.75%) | 3220; 256 (7.95%) |
| O | 1019; 66 (6.48%) | 1499; 93 (6.20%) | 1336; 85 (6.36%) | 2885; 133 (4.61%) | 2859; 213 (7.45%) | 5656; 420 (7.43%) | 15254; 1010 (6.62%) |
| AB | 101; 9 (8.91%) | 73; 6 (8.22%) | 73; 4 (5.48%) | 191; 15 (7.85%) | 214; 18 (8.41%) | 373; 49 (13.14%) | 1025; 101 (9.85%) |
| Rhesus type : |  |  |  |  |  |  |  |
| Rh positive | 1846; 139 (7.53%) | 2657; 192 (7.23%) | 2380; 155 (6.51%) | 5043; 274 (5.43%) | 5051; 395 (7.82%) | 9851; 812 (8.24%) | 26828; 1967 (7.33%) |
| Rh negative | 386; 29 (7.51%) | 584; 42 (7.19%) | 571; 26 (4.55%) | 966; 47 (4.87%) | 1105; 86 (7.78%) | 2165; 174 (8.04%) | 5777; 404 (6.99%) |
| Kell type : |  |  |  |  |  |  |  |
| Kell positive | 183; 18 (9.84%) | 260; 17 (6.54%) | 223; 8 (3.59%) | 489; 27 (5.52%) | 487; 28 (5.75%) | 951; 64 (6.73%) | 2593; 162 (6.25%) |
| Kell negative | 2049; 150 (7.32%) | 2981; 217 (7.28%) | 2728; 173 (6.34%) | 5520; 294 (5.33%) | 5669; 453 (7.99%) | 11065; 922 (8.33%) | 30012; 2209 (7.36%) |

**Figure 1:**
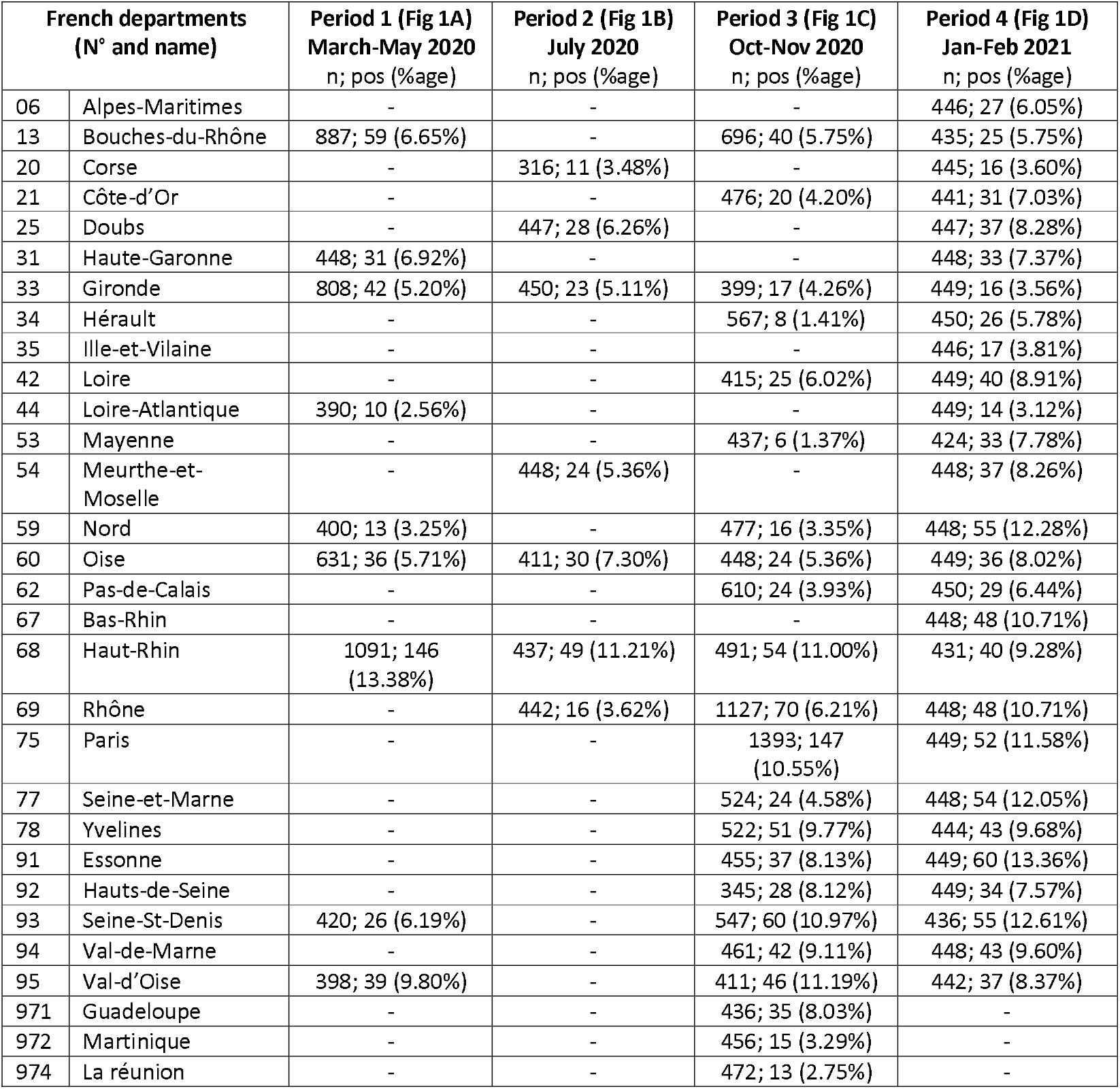
Anti-SARS-CoV IgG (anti-S1) seroprevalence in French departments tested at different time of the epidemics. Fig 1A: samples collected between March and May 2020; Fig 1B: samples collected in July 2020; Fig 1C: samples collected in October and November 2020 and Fig 1D: samples collected in January-February 2021. Numbers labelled on the maps referred to French departments, name of departments, number of samples tested, number of positive samples and percentage of positive sample are displayed in the table below.

**Figure 1:**
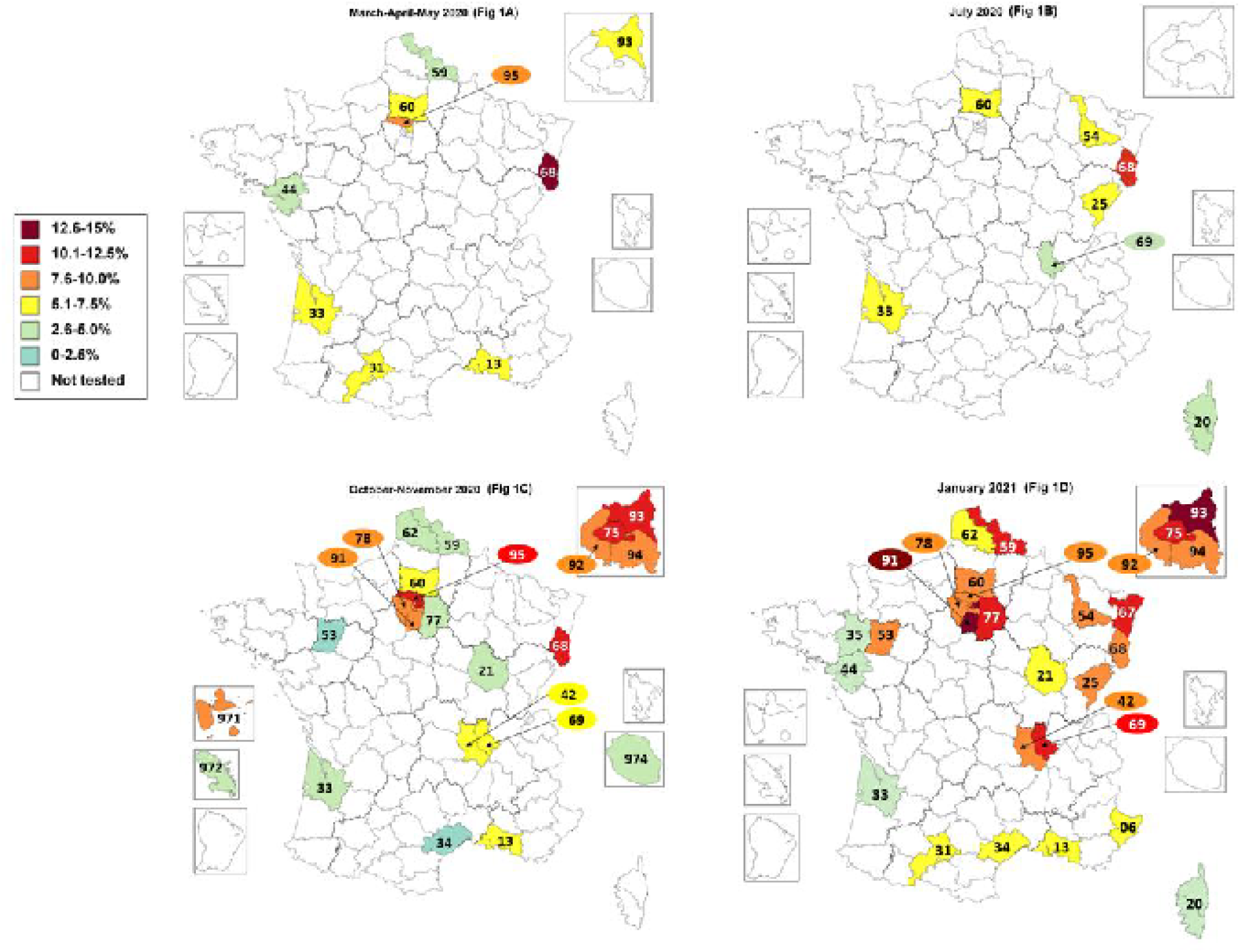

**Figure 2:**
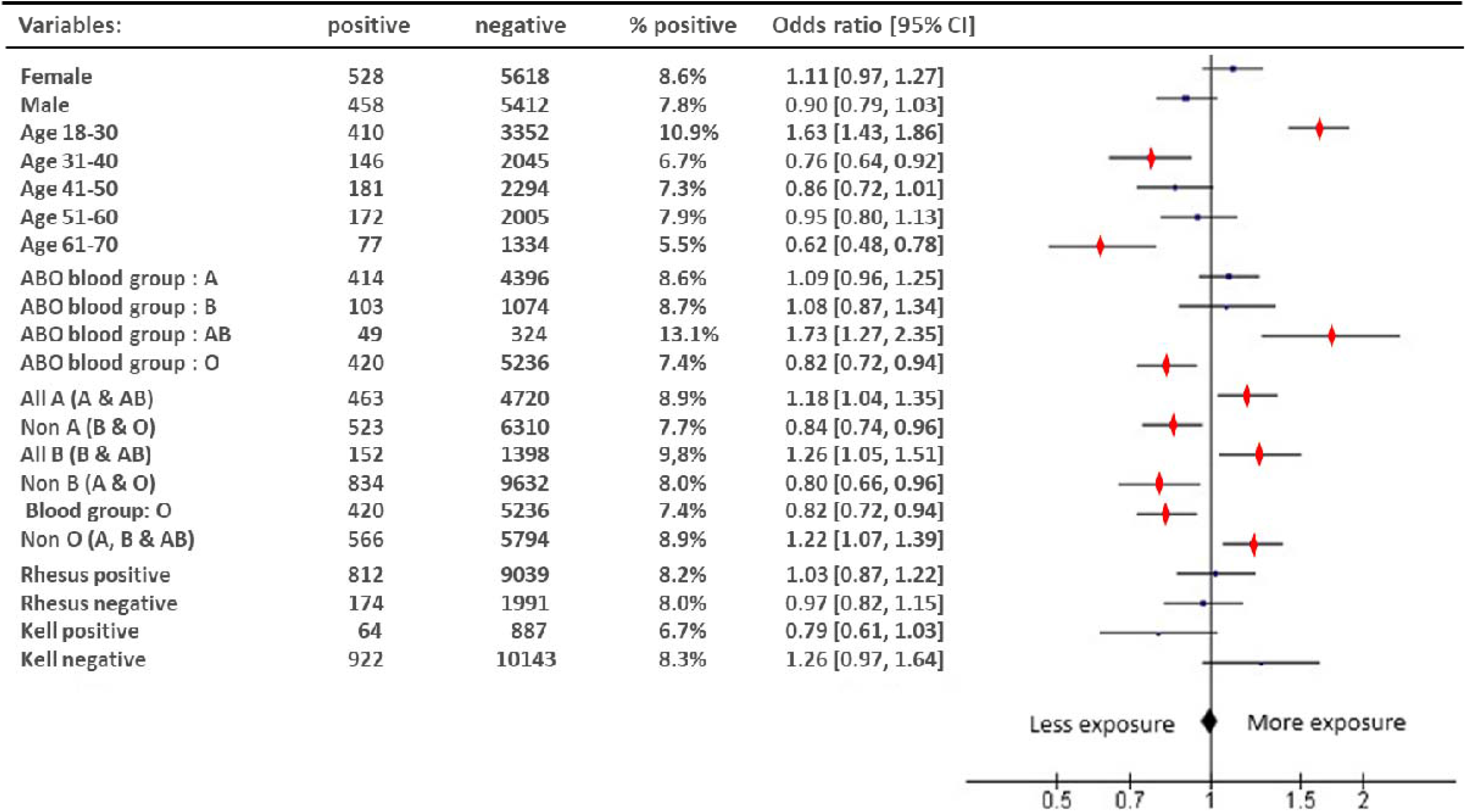
Last survey, January/February 2021 (T6). Forest plot of odds ratios and 95% confident interval reporting association between positive IgG anti-SARS-CoV-2 and blood donation characteristics. In red significant difference, p<0.05.

**Figure 3:**
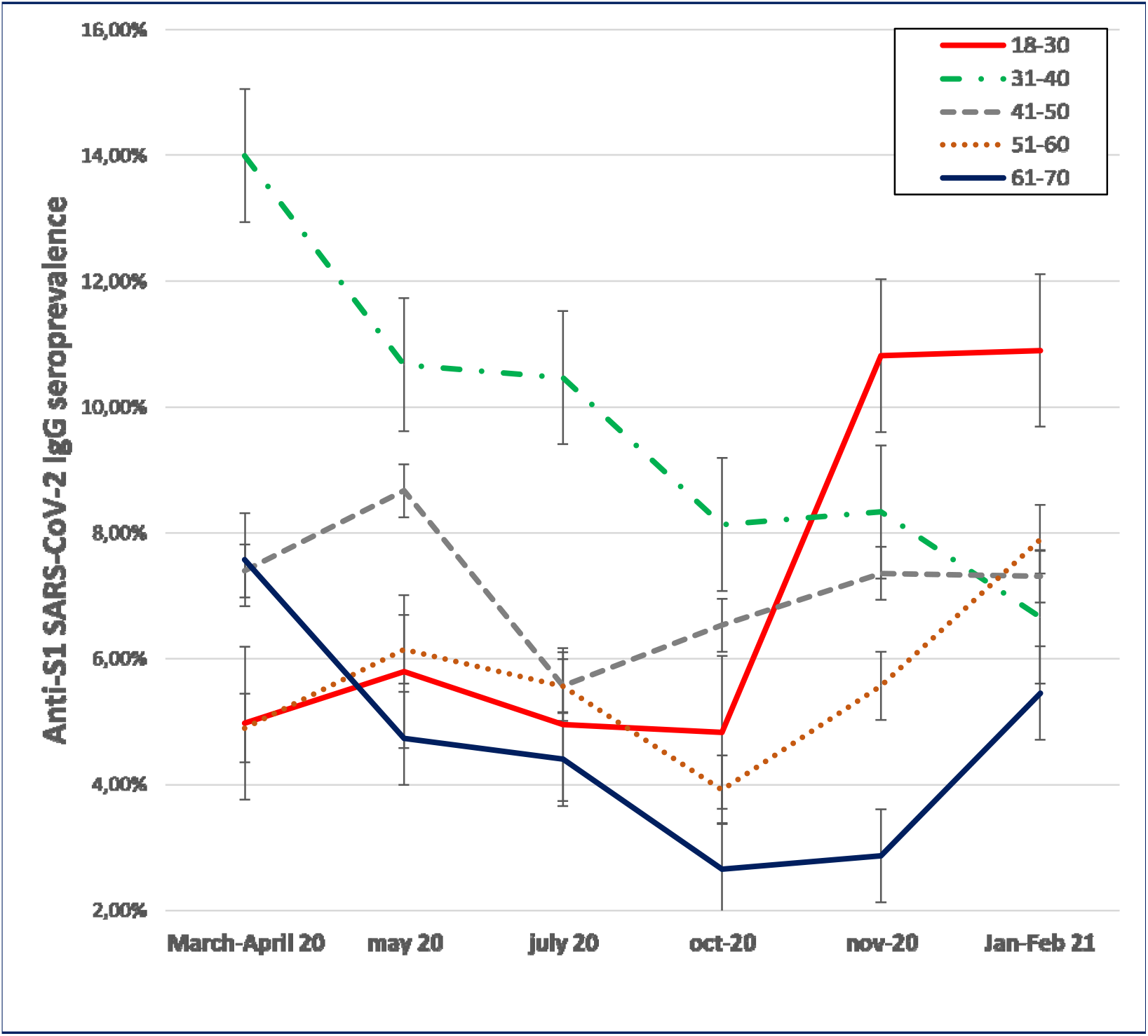
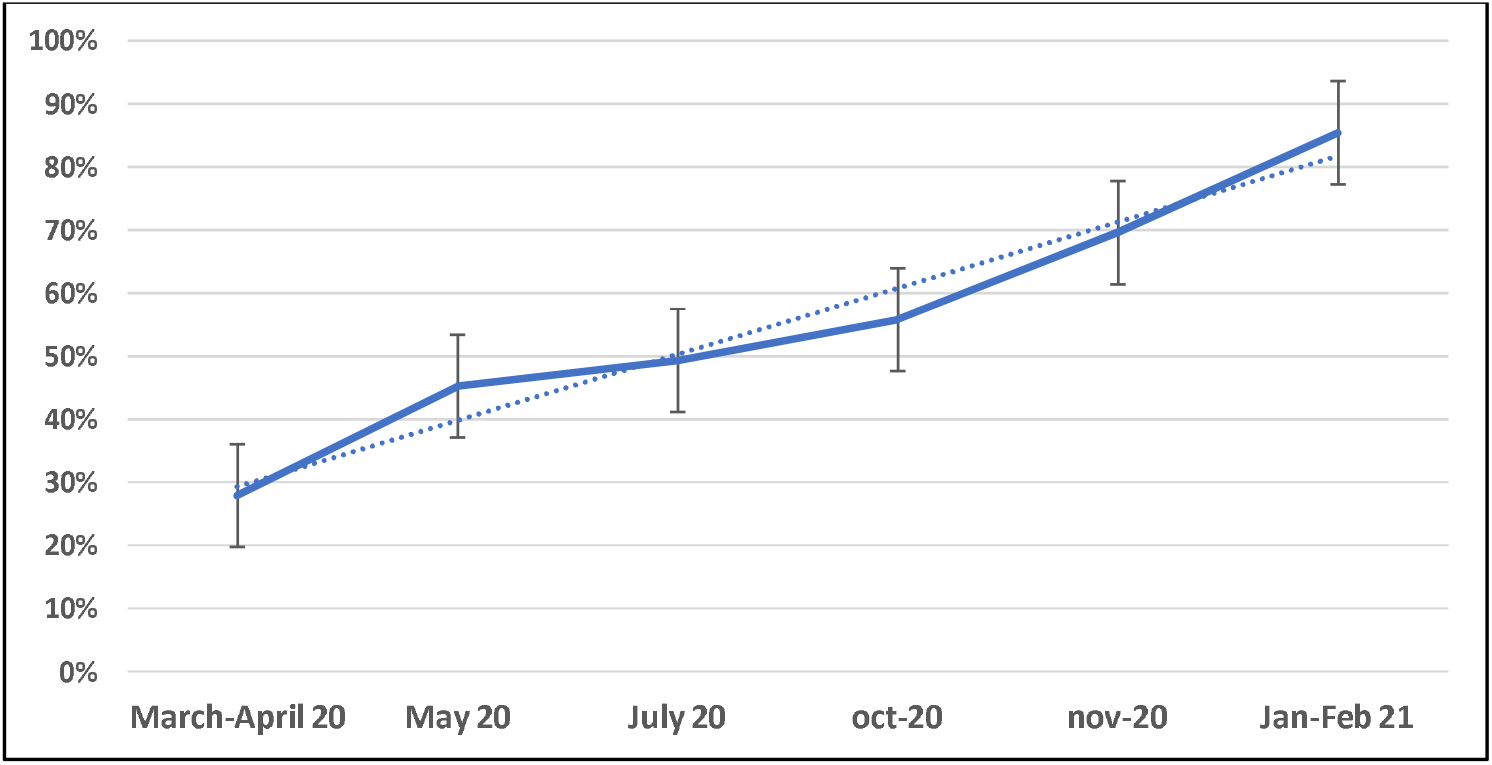
Follow-up of serological markers during the studied period. **A:** Distribution of Anti-S1 SARS-CoV-2 IgG seroprevalence in age groups during the survey. **B:** Proportion of neutralizing antibodies (titers≥20) in positive samples. Linear regression (dotted line) was calculated as y = 0,1048x + 0,1885 and coefficient of regression was R^2^ = 0,9631.

We assessed how IgG seroprevalence rates varied with the phenotypic expression of ABO blood group antigens in donations at the six surveillance periods (Table 1). At T6, of all donations tested (n= 12,016), the prevalence of anti-SARS-CoV-2 IgG in group O donors (7.43%; 95%CI 6.74-8.11) whose red blood cells do not carry A or B antigens was significantly lower (p=0.003) than in groups A, B, and AB donors (8.90%; 95%CI 8.20-9.60). The seroprevalence in groups A and AB (“all A” antigen-carrying) donors was higher (p=0.011) than in “non-A” donors (blood groups B and O): 8.93% (95%CI 8.15-9.71) versus 7.65% (95%CI 7.02-8.28). A similar difference (p=0.014) was observed for “all B” groups: 9.81% (95%CI 8.33-11.29) versus 7.97% (95%CI 7.45-8.49) for “non-B”. No significant difference was observed for the prevalence of anti-SARS-CoV-2 IgG according to Rhesus or Kell blood groups (Figure 2).

The proportion of neutralizing antibodies tested positive (titre >10) in reactive samples (positive and indeterminate) increased during the studied period from 27.9% in March-April 2020 to 85.4% in early 2021 (Figure 3B).

### Model of the dynamics of anti-SARS-CoV-2 IgG prevalence

We developed a mathematical model that combined the anti-SARS-CoV-2 IgG seroprevalence with the daily number of hospital admissions to estimate the probability of hospitalization upon infection and determine the number of infections while correcting for antibody decay. Using our mathematical model, we found that the proportion of population infected during the first wave (March-May 2020) was heterogeneous across France, consistent with previous studies (Hozé et al., 2021; Warszawski et al., 2020), with seroprevalence ranging from 2.9% in Pays de la Loire to 15.8% in Grand-Est region (Figure 4A). The cumulative number of infected individuals remained stable during the summer and increased during the second wave that started in September 2020 (Figure 4B). In contrast, the seroprevalence was stable during the different sampling periods in most regions and even decreased in Grand-Est. In the model, we explained this difference between the number of infected and the seroprevalence by a decay of the antibody following seroconversion.

**Figure 4:**
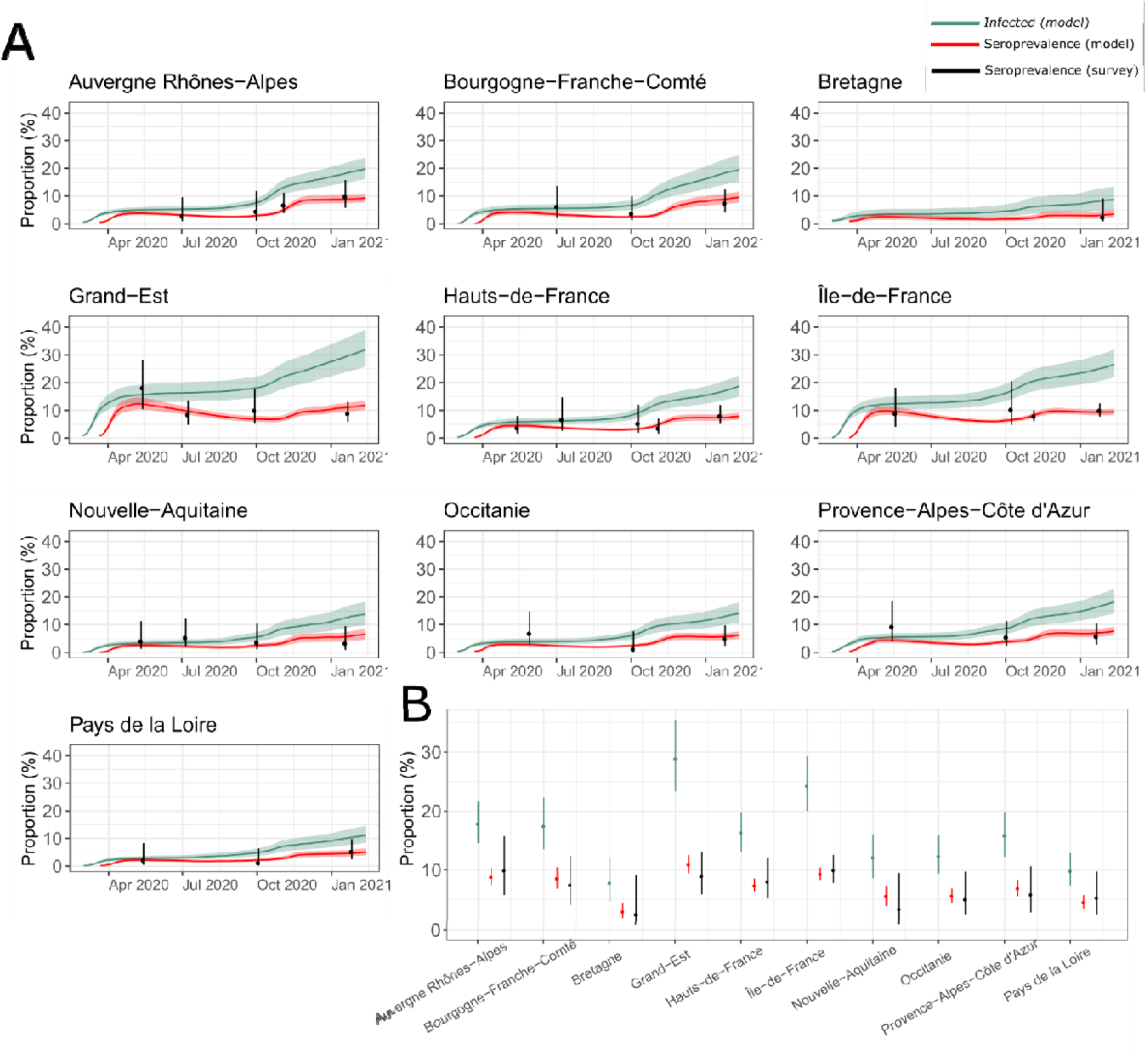
Model-based reconstruction of the cumulative proportion of adults infected. **(A)** Proportion of the population infected by SARS-CoV-2 (green) and reconstructed seroprevalence (red) in ten out of thirteen regions of metropolitan France. The solid lines and envelopes represent the mean and 95% credible intervals, respectively. The dots and bars are the mean and 95% confidence intervals of the observed seroprevalence, aggregated for the different age groups and departments within a region. **(B)** Comparison of the proportion of the adult population infected on Jan, 15, 2021 (green), the corresponding model-based estimate of the seroprevalence (red) and the seroprevalence in the latest sampling January-February 2021 (black). For each administrative region (in italic), the following departments were included in the serostudy: Auvergne-Rhônes-Alpes: *Loire and Rhône* ; Bourgogne-Franche-Comté: *Côte d’Or and Doubs;* Bretagne: *Ille-et-Vilaine* ; Grand-Est: *Meurthe-et-Moselle, Bas-Rhin and Haut-Rhin* ; Hauts-de-France : *Nord, Oise and Picardie;* Ile-de-France : *Paris, Seine-et-Marne, Yvelines, Essonne, Hauts-de-Seine, Seine-Saint-Denis, Val-de-Marne, Val-d’Oise;* Nouvelle-Aquitaine: *Gironde;* Occitanie : *Haute-Garonne and Hérault*, Provence-Alpes-Côte d’Azur: *Bouches-du-Rhône and Alpes-Maritimes*, Pays de la Loire: *Loire-Atlantique and Mayenne*.

We estimated that the capacity of the assay to detect a past infection is 50% of its initial capacity after 4.0 months following seroconversion (95% CrI: 3.0 – 5.2 months) (Figure 5A). As a result, our model predicts that 87% (95% CrI: 78-93) of the individuals infected during the first wave of March-May 2020 were already seronegative by the time of the last sampling (January 2021). This fast antibody decay leads to important differences between the proportion infected and the seroprevalence (Figure 5B), with an estimated proportion of infected individuals in the population being more than two-fold higher than the observed seroprevalence (Figure 5B and Table S1). The infection hospitalization ratio increased with age, from 0.56% (95% CrI: 0.45-0.68) for the 18-30 age group, to 6.75% (95% CrI: 5.19-8.81) in the 61-70 age group (Figure 5C and Table S2). The patterns of infection-hospitalization ratio (IHR) were consistent with those derived from another seroprevalence survey that was conducted in three regions of metropolitan France in May 2020 (Figure 5C) (Carrat et al., 2021; Hozé et al., 2021; Lapidus et al., 2021).

**Figure 5.**
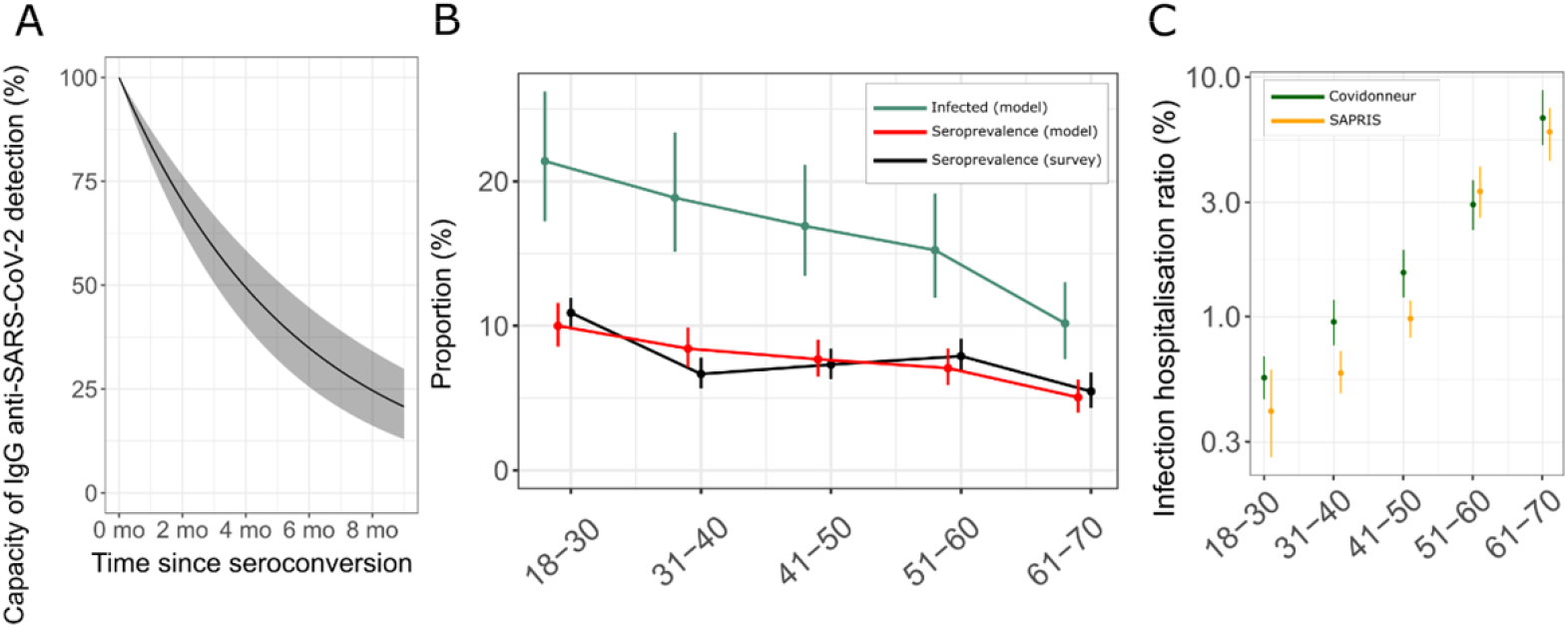
Estimates of the model parameters. **(A)** Capacity of detection of the EuroImmun IgG assay as a function of time since seroconversion. In the model we assume a 100% sensitivity at the time of seroconversion (21 days after infection). **(B)** Comparison of the estimated cumulative number of infections (green), the seroprevalence predicted by the model (red) and observed seroprevalence (black) for the different age groups at the final sampling time (January 2021). The dots and bars are the mean and 95% credible interval for the model-estimated exposure and seroprevalence, and the mean and 95% binomial confidence interval for the observed seroprevalence, respectively. **(C)** Model estimates of the infection-hospitalization ratio. The dots and bars represent the mean and the 95% credible interval of the posterior distribution (in green). We also represent the IHR obtained from a different serological survey (Sapris study, in orange).

The reconstructed seroprevalence data agreed well with the observations (Figure 4A), with 85% (145/170) of reconstructed values falling in the 95% confidence interval of the observations (in the surveys taken across age group, sampling periods, and regions). In an additional sensitivity analysis, we assumed that the decay rate was different for those aged below and above 50 yo and found no difference between the different age groups (Figure S2).

## DISCUSSION

Our objective was to follow the anti-SARS-CoV-2 IgG prevalence rates at the onset of the outbreak in France and to estimate the seroprevalence before the roll out of vaccines. A total of six sero-epidemiological surveys were conducted from March 2020 to January 2021 using blood donor samples (n = 32,605) collected in several French departments.

We investigated seroprevalence at different time points and we deliberately included areas with high incidence. At the time of the first epidemic peak (March-April 2020), the seroprevalence rates observed in France among blood donors were as high as 10.6% (95%CI 8.30-12.90) (Table S3). Seroprevalence rates observed in blood donors were consistent with the number of cases observed in the general population, which was our original criterion for the selection of the departments to be investigated. These data were close to those observed in the Netherlands at the same period, which showed a strong geographical disparity with a maximum of 9.5% in the South of the country (Slot et al., 2020). Such disparity has been reported in many blood donor populations regardless of the study period and the serological tests used and can be explained by the history of the introduction of the virus in the different territories studied, the chronology of implementation and the nature of the control measures decided by the health authorities (Fischer et al., 2020; Jones et al., 2021; Kaspersen et al., 2022; Slot et al., 2020; Stone et al., 2022; Weidner et al., 2021). In the early 2020 period, prevalence reported in Europe were well correlated with case data observed in the regions studied, both in areas of low (eg, Germany, South-East Italy, Denmark, Austria: 0.9% to 2%) or high (eg, Lombardy in Italy: 19.7%) circulation (Cassaniti et al., 2021; Di Stefano et al., 2021; Fischer et al., 2020; Kaspersen et al., 2022; Weidner et al., 2021). In January 2021, at the start of the vaccination campaign, seroprevalence in metropolitan France ranged from 3.1% (95%CI 1.50-4.70) to 13.5% (95%CI 10.34-16.66) depending on the region (Table S3), comparable to that observed in other European blood donor populations: 5.4% to 15% (Castro Dopico et al., 2021; Valenti et al., 2021; Weidner et al., 2021), again with regional disparities (Figure 1).

Monitoring of serological results shows an increase in the proportion of neutralizing antibodies among IgG positive individuals over time (Figure 3B). This may reflect a maturation of the neutralizing response over time and deserves further immunological characterization.

We also investigated the relationship between seroprevalence and blood groups. Previous studies have reported an association between ABO blood types and the risk of exposure to SARS-CoV-2 or the severity of COVID-19: individuals of group O appear to have a lower susceptibility to infection (Cai, 2020; Cheng et al., 2005; Dai, 2020), while those of group A (and in some studies those of groups B and/or AB) are more likely to be infected (Pendu et al., 2021). In a previous study, we observed a lower seroprevalence of neutralizing anti-SARS-CoV-2 antibodies in blood donors of group O compared to those of other groups (A, B, and AB) (Gallian et al., 2020). These preliminary data are confirmed in the population of donors collected in 2021 (n=12,016). The proportion of donors carrying anti-SARS-CoV-2 IgG from group O (7.43%; 95%CI 6.74-8.11) is lower (p=0.003) than that of the other groups (8.90%; 95%CI 8.20-9.60) with an odds ratio of 0.82 [0.72-0.94] (Figure 2). Our results are in agreement with data reported in other blood donor surveys (Butler et al., 2022; Vassallo et al., 2021; Valenti et al., 2021). One previously proposed hypothesis to explain the protective effect observed in blood group O individuals is that the natural anti-A and anti-B antibodies present in group O individuals may limit the interaction between the viral spike protein and the ACE-2 receptor and thus play a role in susceptibility to infection (Deleers et al., 2021; Zhao et al., 2021).

In addition, we investigated seroprevalence by age group. Among older donors, lower anti-SARS-CoV-2 IgG seroprevalence is frequently reported in the literature (Dodd et al., 2020; Jones et al., 2021; Stone et al., 2022; Valenti et al., 2021; Weidner et al., 2021). In our study, the age group 61-70 years (Figure 3A) had the lowest prevalence throughout the study, except during the first epidemic wave. In the first samples (March-April 2020), the presence of antibodies reflected early exposure to SARS-CoV-2, before the implementation of specific protective measures, including a strict lockdown between 17 March and 11 May 2021. Subsequently, lower prevalence rates among the elderly reflected adherence of this group to prevention measures implemented to protect populations most vulnerable to COVID-19. In those under 30 year old, we observed an increase in seroprevalence in the autumn of 2020, reflecting increased exposure during the summer and early autumn. This increase, probably linked to a relaxation of preventive measures during the summer, occurred in a context of low detection of incident cases due to the frequency of a- or pauci-symptomatic forms in the youngest age group.

We extended our previous approach which used a mathematical model (Hozé et al., 2021), to reconstruct the number of infections from hospital surveillance data and serological data. This latter approach was based on a serological study from May 2020 where the waning of immunity was not observable. Here we accounted for the decay in the ability to detect IgG anti-S1 antibody by the serological assay used in our study. Sampling for the serological study performed at different time points – and during time period long enough – allowed us assessing the antibody decay. Our approach is similar to that of Chen *et al*. who reconstructed the number of infections using serological surveys in blood donors across England and used mortality data to estimate the infection fatality rate (Chen et al., 2021). However, since mortality is low in the younger age groups we used hospitalization data. We assumed that probability of hospitalization was stable during the period under review. Relaxing this assumption may lead to different parameter estimates.

Our analysis provided the proportion of infected adults by region and age group. Hospital admissions vary greatly between departments, partly because individuals may go to hospitals in the neighboring departments. In the hospitalization SI-VIC database, we were able to access the department where individuals are admitted to hospital but not their place of residence. By aggregating by region and not by department, we reduced this bias. We reported a very high rate of infections in the Grand-Est region, but the serological surveys were mainly performed in the Haut-Rhin department, which may not be representative of the virus circulation in the whole region. Otherwise, our data were consistent with the predictions made from SAPRIS, another large-scale serosurvey in the general population (Figure S3) (Hozé et al., 2021). The lower proportion of infected individuals in the SAPRIS survey can be explained by the fact that all adults were included, including those aged 70 and over who had a lower infection rate.

Overall, the use of this model has shown a high degree of consistency between the hospitalization data observed at a given time in the general population and the results of seroprevalence studies in blood donors conducted several weeks later. A similar finding was reported in a seroprevalence study of SARS-CoV-2 in adult donors, where the results were comparable to those obtained in household surveys targeting the general population (Grant et al., 2021). Another advantage of the model is that it is possible to estimate IgG seroprevalence rates from hospitalization data for territories where serological surveys have not been conducted. The use of this model therefore makes it possible to provide health authorities with rapid information on the proportion of the adult population that has been in contact with the virus.

The impact of seroreversion or loss of IgG detection capacity is difficult to consider at any given time. Indeed, regions have been diversely affected with varying chronology of viral circulation and intensity over time. However, the model is able to mitigate these variations and provides an estimate of the decay time of IgG-S1 antibody detection by the Euroimmun test. The modelling data show that approximately 50% of individuals who are IgG-S1 positive have no detectable antibodies four to five months after infection (Figure 5A). In a study similar to ours conducted before the vaccination campaign in Italy (July 2020 to February 2021), the authors report that about 50% of anti-RBD IgG positive individuals became negative after a median follow-up of 18 weeks (4.5 months) (Valenti et al., 2021).

In conclusion, in this study, we showcased insights we could get from SARS-CoV-2 seroprevalence studies performed in blood donors in France. We observed differences in anti-SARS-CoV-2 IgG seroprevalence according to geographical distribution and over time. The seroprevalence in France before the start of the vaccination campaign was about 15%. The study showed that older people, who had the lowest seroprevalence rates, had complied with prevention measures. It identified an increase in exposure in the under-30s during the summer of 2020 that had gone unnoticed due to the high proportion of asymptomatic or pauci-symptomatic infections in this population. The study of blood group distribution in IgG-S1 positive individuals showed a slightly lower exposure to SARS-CoV-2 infection in group O donors compared to non-O donors. Over the course of the study, the proportion of neutralizing antibodies in IgG-positive samples increased, suggesting a maturation of the immune response. The use of a mathematical model showing a high degree of consistency in the different regions studied between general population hospitalization data and the results of seroprevalence studies in blood donors allows for rapid and extemporaneous quality estimates of the level of immunity acquired by the general population to be provided to health authorities.

### Limitations of the study

Our study has several limitations: (i) the geographical areas investigated were chosen as representative of regions with different intensities of viral circulation and the study was not performed for the whole territory. Although the magnitude and dynamics of prevalence in each area are robust and of the same order of magnitude as data observed in other European countries, the study was not designed to provide an accurate overall seroprevalence. (ii) The blood donor population does not allow exploration of seroprevalence in the <18 and >70 age groups. (iii) Blood pre-donation selection measures exclude individuals with recent clinical signs suggestive of COVID-19 and those with persistent sequelae that may lead to an underestimation of rates. (iv) Individuals may be treated in hospitals in neighboring departments. In the SIVIC database, we have the department where individuals are admitted to the hospital but not their place of residence. In our study, the mathematical model allows us to determine the proportion of infected adults by region and age group. Aggregating by region and not by department reduced this bias.

## Data Availability

The paper used aggregated data and individual participant data will not be made available. Data are provided in a format that maintains anonymity of the participants. For each individual, we provide the age group (in groups 18-30, 31-40, 41-50, 51-60, 61-70), department, survey period and median date of the survey. Code and data are available on https://gitlab.pasteur.fr/mmmi-pasteur/covidonneur

## ACKNOWLEDGMENTS

We are grateful to blood donors who participated in the study. We thank the regional French National Blood Service directors and their field teams for their highly contributive participation for blood collection: ETS Auvergne-Rhône-Alpes : Dr D. Legrand, Dr J. Courchelle, Dr V. Barlet; ETS Bourgogne-Franche-Comté : Dr C. Besiers; Dr C. Barisien; ETS Bretagne : Dr B. Danic, Dr A. Guillard; ETS Centre-Pays-de-la-Loire : Dr. F. Bigey, Dr C. Leclerc, Dr S. Le Cam; ETS Grand Est : Dr C. Gachet, Dr C. Claudel; ETS Guadeloupe-Guyane & ETS Martinique : Dr F. Maire, Dr M. Bordenet, Dr D. Arnaudova; ETS Hauts-de-France-Normandie : Dr R. Courbil, Dr N. Delemer, Dr C Narboux; ETS Ile-de-France : Mr S. Noel, Dr A. Slimani; ETS Nouvelle-Aquitaine : Dr M. Jeanne, Dr F. Lassurguere; ETS Occitanie : Dr L. Bardiaux; Dr P. Lambert, Dr C. Maugard; ETS Provence-Alpes-Côte-d ‘Azur-Corse: Prof. J. Chiaroni, Dr. C. Lazaygues; ETS La-Réunion-Océan-Indien : Dr I. Delouane, A. Le Tallec. We thank B. Bonneaudeau, A. Michaut and K. Bornarel for their support and expert advice concerning ethic and juridic aspects.

This study was performed with the financial support of the Agence Nationale de la Recherche (ANR-20-COVI-0073-01, France), the Fondation Recherche Médicale and the ANRS Maladie Infectieuses Émergentes (ANRS-MIE). SC acknowledges financial support from the Investissement d’Avenir program, the Laboratoire d’Excellence Integrative Biology of Emerging Infectious Diseases program (grant ANR-10-LABX-62-IBEID); the INCEPTION project (grant PIA/ANR-16-CONV-0005); the European Union’s Horizon 2020 research and innovation program (grants RECOVER 101003589 and VEO 874735); AXA; and Groupama.

## AUTHORS CONTRIBUTIONS

XdL, PM and PG Designed the study. NB, CI, PMSV, EN, BP performed the sample acquisition, laboratory testing (Elisa and/or seroneutralization assays) and data analysis. PG, XdL interpreted the virological findings. NH and SC made and interpreted the modelling studies. PG, NH drafted the manuscript. PM, PR, XdL and SC reviewed the manuscript. All co-authors contributed to the editing and final drafting of the manuscript and figures. All authors approve the content and submission of the paper.

## DECLARATION OF INTERESTS

PG, NB, CI, PR and PM are employed by the French transfusion public service (Etablissement Français du Sang) in charge of blood products manufacturing and issuing in France. The funders had no role in the design of the study; in the collection, analyses, or interpretation of data; in the writing of the manuscript, or in the decision to publish the results.

## METHODS

### Study design and population

This is a prospective repeated cross-sectional study for -SARS-CoV-2 anti-S1 IgG prevalence in French blood donor population. Sampling was performed at six different times (T1= end March–early April, T2= May, T3 =July, T4= October, T5= November 2020 and T6= mid-January–early February 2021) during the epidemic period prior to the implementation of the vaccination campaign and the circulation of antigenic variants (Figure S1).

Voluntary unpaid blood donors (18-70 years old) were recruited according to French regulatory requirements. The pre-donation medical questionnaire included the reporting of influenza-like clinical symptoms. During the COVID-19 epidemic, donors reporting a history of clinical and/or biological diagnosis or suspicion of SARS-CoV-2 infection were deferred for 28 days after recovery.

Our study was carried out on donors living in mainland France and in three overseas administrative areas: Guadeloupe and Martinique in the Americas and La Réunion in South-West Indian Ocean. The French territory is divided into administrative areas called ‘departments’, corresponding to NUTS3 level (96 on the mainland and five overseas). Geographical distribution of Anti-SARS-CoV-2 seropositivity was assessed at the departmental level. When reconstructing the dynamics of infection with a model, we aggregated the serological data at the regional level (NUTS1 level).

The evolution of the epidemic was systematically monitored in three departments, two of which were selected because significant viral circulation was observed at the start of the epidemic (Oise (60) in Île-de-France and Haut Rhin (68) in eastern France) and one with low viral circulation (Gironde (33)) in South West France. In order to allow for a follow-up, during the last epidemiological survey (T6: January/February 2021), most departments previously studied in mainland France (n=24) were tested.

Donors recruited in the study were assigned to one of five age groups (18–30, 31–40, 41–50, 51–60, and 61–70). The information made available for analysis included date of donation, sex, age, and results for ABO, Rhesus, and Kell blood phenotyping.

### Ethical statement

Only volunteer blood donors were included. They were informed that samples would be tested for blood-borne pathogens in order to prevent transfusion-transmitted infections and might be used for epidemiological studies and more specifically for the present study. The study was approved by the scientific direction of the EFS and an ethic committee (CPP Ouest 6-CPP 1286/MS1, N°IDRCB: 2020-A01200-39). All data used for epidemiological studies were de-identified.

### Laboratory methods

#### Anti-SARS-CoV-2 IgG detection

ELISA detection of SARS-CoV-2 specific IgG was performed using the ELISA SARS-CoV-2 IgG assay (Euroimmun, Lubeck, Germany) targeting the S1 sub-unit of the spike protein that include the RBD (receptor binding domain) region. Serology tests were performed according to manufacturer’s instruction. For each sample, we calculated the ratio sample OD/cut-off OD. A result was considered positive if the sample ratio was⍰≥⍰ 1.1.

#### Anti-SARS-CoV-2 neutralizing antibodies titration

All reactive samples with ELISA positive or indeterminate results were tested using a virus neutralization test (VNT) as previously described for titration of anti-SARS-CoV-2 neutralizing antibodies (Gallian et al., 2020). All samples were tested in duplicate for all dilution (1:10 to 1:160) and considered positive for titre above 10 using a D614G B.1 European strain (BetaCoV/Germany/BavPat1/2020, courtesy of Pr C Drosten).

#### lood grouping

ABO/Rh/Kell phenotypes were determined using a fully automated microplate haemagglutination procedure according to routine EFS procedures.

### Statistical analysis

Association of the presence of anti-SARS-CoV-2 IgG antibodies with biological factors (Blood groups) and demographic data (sex, age, place of residence) was analyzed using the Chi-square test (“Chi-Square Test Calculator” http://www.socscistatistics.com/tests/chisquare2/Default2.aspx). Results were considered statistically significant when p-value was lower than 0.05. Univariate and multivariate analysis were performed using the IBM-SPSS Statistics v 24.0.0.0 software (Chicago, IL, USA).

### Hospitalization data

The number of hospitalized patients was obtained from the SI-VIC database (système d’information pour le suivi des victimes - patient monitoring information system), used to monitor hospitalizations in the event of exceptional health situations. This database is maintained by the French ANS (Agence du Numérique en Santé) and provides real time information on COVID-19 patients hospitalized in public and private French hospitals. Data, including age, hospitalization date, outcome and region, are sent daily to Santé Publique France, the French national public health agency. All COVID-19 cases registered are either biologically confirmed or present with a computed tomographic image highly suggestive of SARS-CoV-2 infection.

### Reconstruction of the dynamics of infection and observed seroprevalence

We designed a statistical framework that combines the observed seroprevalence at different time points with a model of antibody decay to retrieve the total number of infections. Using the number of hospital admissions, we reconstructed the dynamics of the infection using a deconvolution approach, assuming an infection-to-hospitalization delay of 11 days. Since only a fraction of the infections result in hospitalization, we introduced a probability of hospitalization to obtain an estimate of the number of infections. The vast majority of infected individuals (with severe as well as asymptomatic forms of the disease) have developed IgG approximately 21 days post infection. We used the cumulative number of infections divided by the total adult population within a region as a measure of the proportion infected. A model of exponential decay of the sensitivity of the assay was then applied to explain the observed seroprevalence at the different time points.

More specifically, we denote *x*_i_ and *n*_i_ the number of seropositive and number of samples within group *i* with shared characteristics (at a given date, age class, location). The infection-to-hospitalization delay distribution is modelled with a gamma distribution (mean = 11 and sd = 3.3). The number of hospitalizations on day *t* is given by the convolution of the delay with incident infections

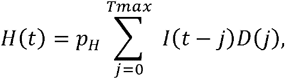

where *P*_*H*_ is the hospitalization probability, *D(j)* is the probability that infection-to-hospitalization delay is *j* days, and *Tmax* is 400 days. This relation is inverted to reconstruct the daily number of infections *I(t)* using the deconvolution procedure as previously described (Hozé et al., 2021).

Since antibody levels decrease over time, we modelled the expected seroprevalence as the cumulative number of infections modulated by the time since seroconversion (conservatively estimated at 21 days post-infection). We assumed an exponential decay of the sensibility of the serological assay. The seroprevalence on day *t* is given by

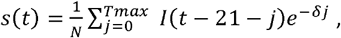

where *N* is the population size. The likelihood function is obtained over all locations, age groups, and sampling periods:

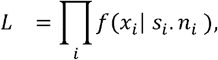

where *s*_*i*_ is the seroprevalence for group i on the day corresponding to the median date of the blood collection for the given sampling period (see Table 1) and *f(·*|*λ)* is a Poisson distribution of mean *λ*. In addition, we assumed regional differences in the probability of hospitalization.

We estimated the parameters *P*_*H*_ for age groups 18-30, 31-40, 41-50, 51-60, 61-70, and the decay. A Markov Chain Monte Carlo (MCMC) approach was used to estimate jointly the probability of hospitalization upon infection and the dynamics of the sensitivity of the antibody assay and was implemented in Rstan (Stan Development Team. RStan: the R interface to Stan. R package version 2.19.3. http://mc-stan.org [2020]).

We run four chains of 10,000 iterations each and remove 50% for burn-in. Flat priors were chosen for all parameters. We used 2.5% and 97.5% percentiles from the resulting posterior distributions for 95% credible intervals (CrI) for the parameters.

## Supplemental Information

### Supplemental Information Figures

**Figure S1 :**
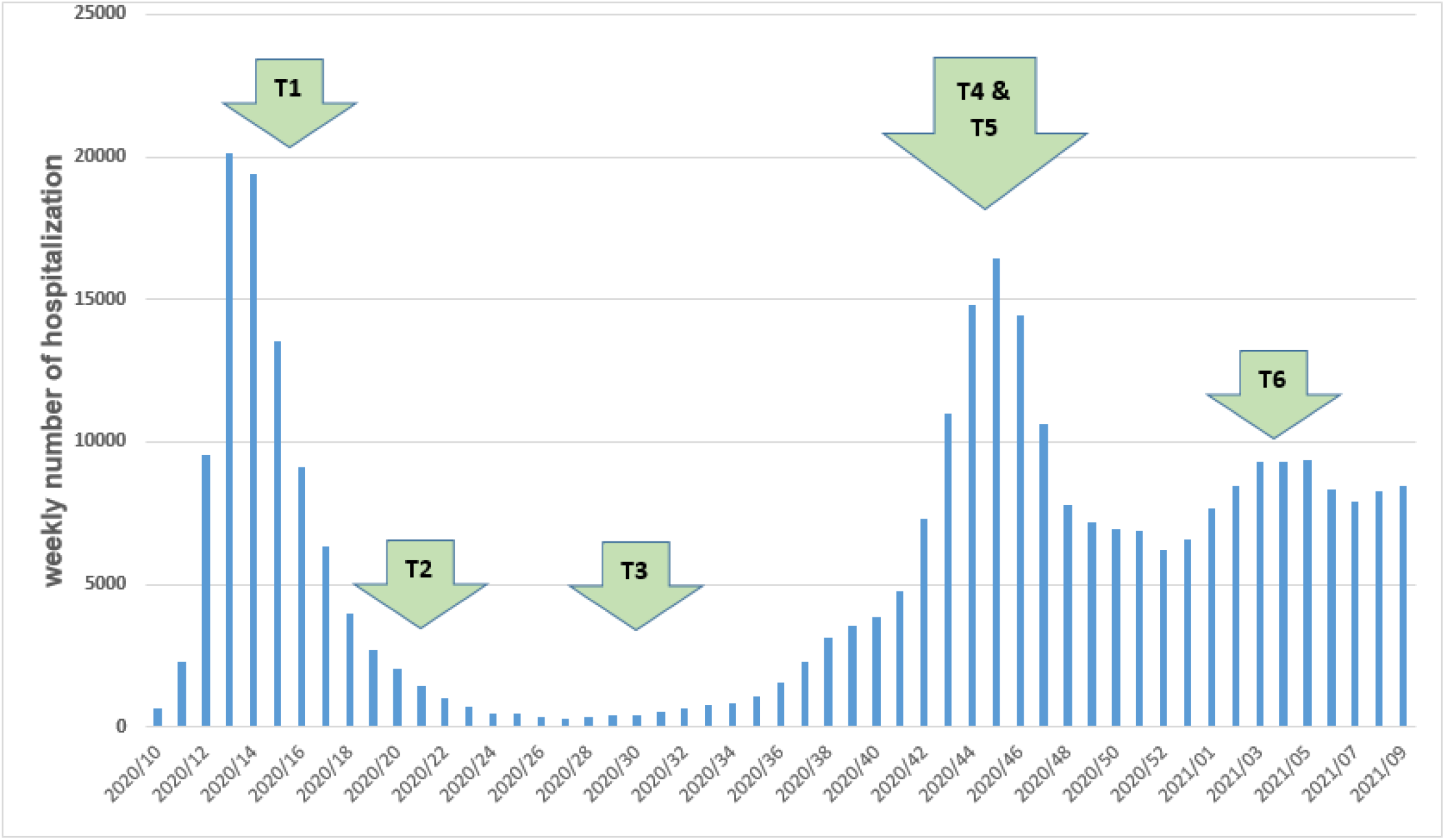
Flowchart of the study according to the epidemiological situation in general population in mainland France. Weekly number of patient admitted in hospital for COVID-19 in France was used as a reliable marker of the epidemiological situation. Supplemental information associated to Table1.

**Figure S2 :**
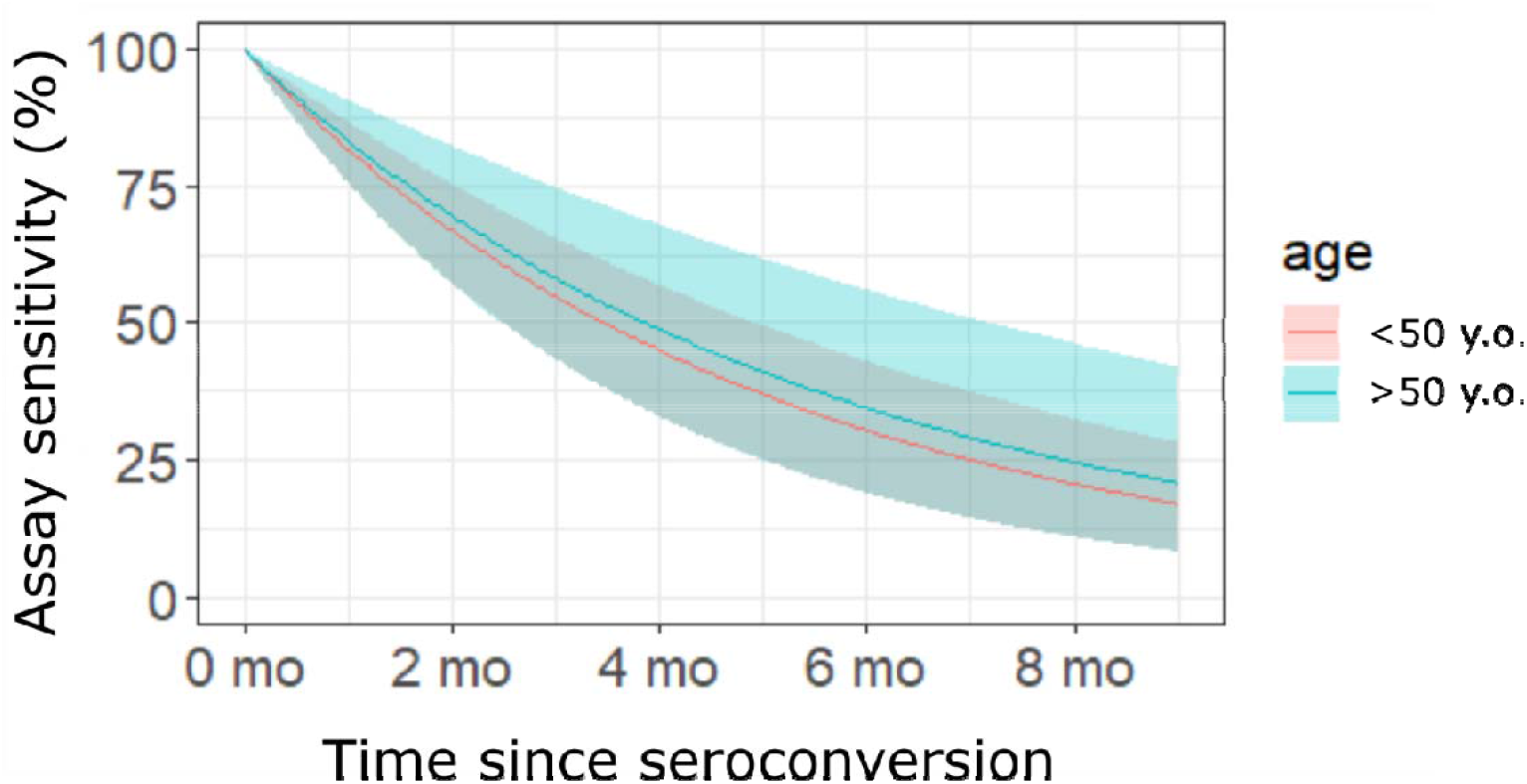
Decay of the assay sensitivity vs time post-seroconversion, when considering a difference below and above 50 year old (yo). Supplemental information related to Figure 5A.

**Figure S3 :**
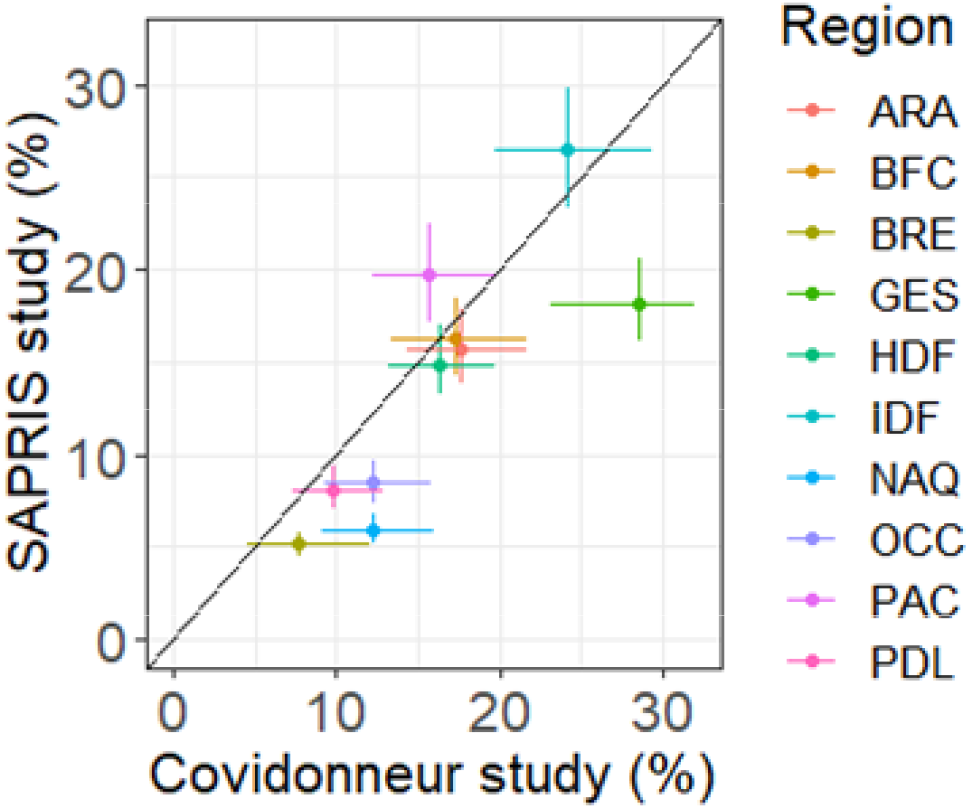
Scatter-plot of the proportion infected in the different regions of metropolitan France, as estimated by the present study (x-axis) and with the Sapris study (y-axis), on January 15, 2021. The dots and bars are the mean and 95% credible interval. ARA=Auvergne-Rhône-Alpes. BFC=Bourgogne-Franche-Comté. BRE=Bretagne. GES=Grand Est. HDF=Hauts-de-France. IDF=Île-de-France. NAQ=Nouvelle-Aquitaine. OCC=Occitanie. PAC=Provence-Alpes-Côte d’Azur. PDL=Pays de la Loire.

### Supplemental Information Tables

**Table S1:** Seroprevalence by age group in January 2021. Seroprevalence from blood donor survey, seroprevalence estimated with the model, and number of infected individuals estimated with the model on January 15th 2021 (data presented in Figure 5B). The number presented are the point estimate and 95% binomial confidence interval for the data, and the mean and 95% credible interval of the posterior distribution for the model estimates.

| Age | Seroprevalence (survey) (%) | Seroprevalence (model) (%) | Infected (model) ( |
| --- | --- | --- | --- |
| 18-30 | 10.9 [9.92 – 11.9] | 9.99 [8.57 - 11.58] | 21.4 [17.23 - 26.2 |
| 31-40 | 6.66 [5.66 – 7.79] | 8.42 [7.13 - 9.88] | 18.86 [15.13 - 23. |
| 41-50 | 7.31 [6.32 – 8.41] | 7.68 [6.48 - 9.04] | 16.9 [13.46 - 21.1. |
| 51-60 | 7.90 [6.80 – 9.11] | 7.07 [5.9 - 8.42] | 15.23 [11.94 - 19. |
| 61-70 | 5.46 [4.33 – 6.77] | 5.05 [3.99 - 6.27] | 10.17 [7.69 - 13.0 |

**Table S2:**
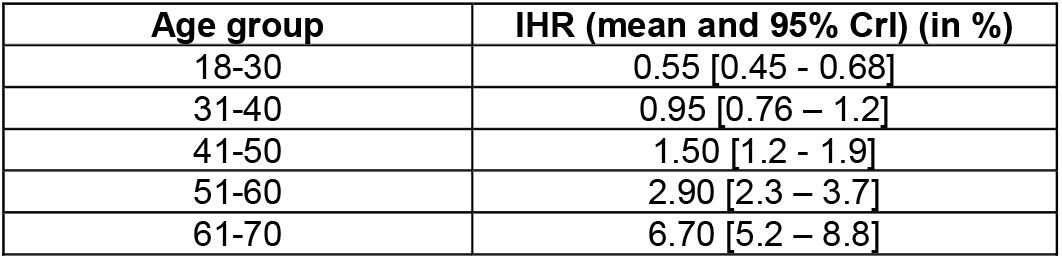
Infection-hospitalization ratio by age group. (data presented in Figure 5C).

| Age group | IHR (mean and 95% CrI) (in %) |
| --- | --- |
| 18-30 | 0.55 [0.45 - 0.68] |
| 31-40 | 0.95 [0.76 – 1.2] |
| 41-50 | 1.50 [1.2 - 1.9] |
| 51-60 | 2.90 [2.3 – 3.7] |
| 61-70 | 6.70 [5.2 – 8.8] |

**Table S3:** Weighted prevalence of IgG antibodies to anti-SARS-CoV-2 after correction for the number of residents and for the age distribution in the general population (26 departments in mainland France).

| Departments | T1 | T2 | T3 | T4 | T5 | T6 |
| --- | --- | --- | --- | --- | --- | --- |
| 13 | 5.6 % | 8.2 % |  | 5.7 % |  | 5.6 % |
| 93 | 6.4 % |  |  |  | 11.1 % | 12.7 % |
| 60 | 7.8 % | 4.6 % | 7.3 % | 5.4 % |  | 8.0% |
| 68 | 10.6 % | 17.9 % | 11.3 % | 11.0% |  | 9.3 % |
| 33 | 5.9 % | 4.6 % | 5.0% | 4.1 % |  | 3.3 % |
| 31 |  | 6.9 % |  |  |  | 6.9 % |
| 59 |  | 3.3 % |  |  | 3.4 % | 12.1 % |
| 95 |  | 10.0% |  |  | 11.6 % | 8.3 % |
| 44 |  | 2.4 % |  |  |  | 3.1 % |
| 54 |  |  | 5.4 % |  |  | 8.4 % |
| 25 |  |  | 6.2 % |  |  | 8.2 % |
| 69 |  |  | 3.6 % | 3.8 % | 7.8 % | 10.7 % |
| 75 |  |  |  | 10.8 % | 10.4 % | 11.6 % |
| 34 |  |  |  | 1.4 % |  | 5.7 % |
| 53 |  |  |  | 1.4 % |  | 7.8 % |
| 21 |  |  |  | 4.1 % |  | 6.6 % |
| 94 |  |  |  |  | 9.1 % | 9.7 % |
| 78 |  |  |  |  | 9.9 % | 9.7 % |
| 77 |  |  |  |  | 4.7 % | 12.1 % |
| 91 |  |  |  |  | 8.4 % | 13.5 % |
| 62 |  |  |  |  | 4.0% | 6.4 % |
| 42 |  |  |  |  | 6.0% | 8.8 % |
| 92 |  |  |  |  | 8.3 % | 7.6 % |
| 67 |  |  |  |  |  | 10.5 % |
| 35 |  |  |  |  |  | 3.6 % |
| 6 |  |  |  |  |  | 5.8 % |

## Notes

### Competing Interest Statement

PG, NB, CI, PR and PM are employed by the French transfusion public service (Etablissement Francais du Sang) in charge of blood products manufacturing and issuing in France. The funders had no role in the design of the study. In the collection, analyses, or interpretation of data; in the writing of the manuscript, or in the decision to publish the results.

### Author Declarations

Only volunteer blood donors were included. They were informed that samples would be tested for blood-borne pathogens in order to prevent transfusion-transmitted infections and might be used for epidemiological studies and more specifically for the present study. The study was approved by the scientific direction of the EFS and an ethic committee (CPP Ouest 6-CPP 1286/MS1, Number IDRCB: 2020-A01200-39). All data used for epidemiological studies were de-identified.

